# Heart Rate Circadian Oscillations as Digital Biomarkers of Cardiometabolic Health Determinants

**DOI:** 10.64898/2026.06.07.26355124

**Authors:** Alessandro Colitta, Simone Bruno, Davide Benedetti, Domeniko Hoxhaj, Francy Cruz-Sanabria, Caterina Di Pede, Francesca Buracchi Torresi, Paolo Frumento, Luna Gargani, Monica Fabbrini, Michelangelo Maestri Tassoni, Enrica Bonanni, Ugo Faraguna

## Abstract

**AIMS:** Cardiometabolic risk factors may impair health by altering the autonomic modulation of the cardiovascular system, a physiological process described by heart rate (HR) circadian oscillations. However, the impact of cardiometabolic health determinants on HR circadian oscillations remains scarcely characterized in real-world, population-based settings. To address this, we applied digital health technologies to investigate how cardiometabolic health determinants shape HR circadian oscillations in a real-world cohort of individuals free of cardiometabolic diseases.

**METHODS:** First, a 10-fold cross-validation of a model was performed, aiming at mitigating wearables’ measurement error caused by motion artifacts. This process was informed by 10,056 epochs of concurrent wearable-derived and polysomnographic HR assessment, yielding an average 1.3 bpm reduction in wearables measurement error. We subsequently applied this model to over 2 million 1-minute epochs of HR data, derived from 7-day continuous actigraphic recordings of 245 individuals free of cardiometabolic disorders. Functional-on-scalar regression modelling and both parametric and nonparametric analyses characterized HR circadian profiles and their relationships with demographics, lifestyle, chronotype, sleep health, and chronic insomnia diagnosis. A 6-dimension sleep health index was calculated.

**RESULTS:** Sex, chronotype, and sleep health predominantly shaped HR circadian oscillations. In detail, females consistently showed higher HR across the 24 hours. Moreover, chronotype was associated to a phase shift in HR circadian profiles, with later timings corresponding to eveningness. Notably, sleep health impacted HR circadian oscillations in a dose-dependent fashion: each additional impaired sleep dimension was associated with a 1.2 bpm HR increase during nighttime, alongside reduced circadian robustness and delayed oscillation timings. Finally, the earlier occurrence of morning HR peaks served as a digital biomarker of insomnia (80% specificity, 74% sensitivity).

**CONCLUSIONS:** This work provides a digital health framework to characterize HR circadian oscillations in free-living populations and supports its clinical utility in capturing the autonomic disruptions related to cardiometabolic health determinants.

**GRAPHICAL ABSTRACT:** 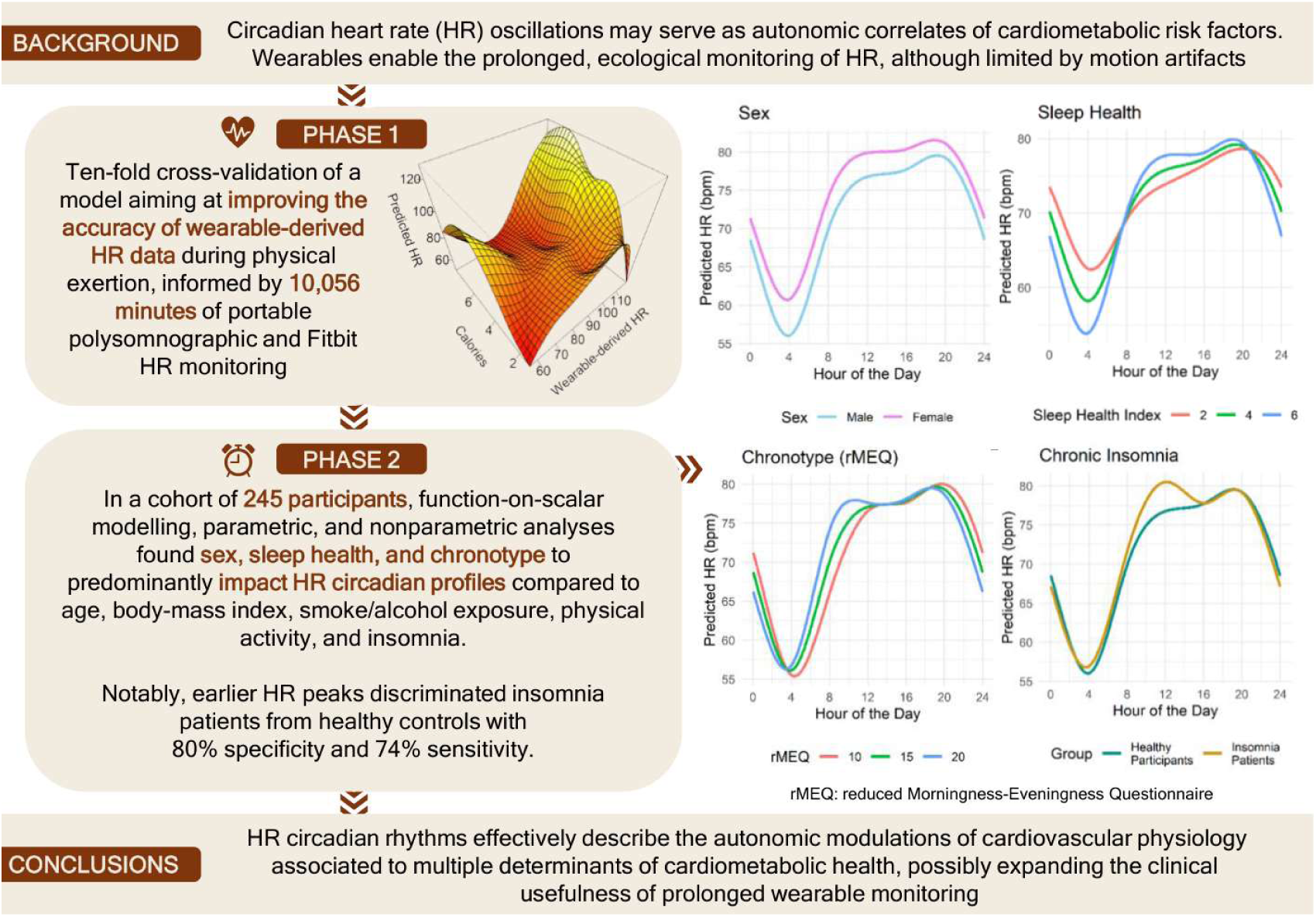

## INTRODUCTION

Wearables are digital health tools allowing for continuous, ecological, and prolonged monitoring of heart rate (HR) ^1^. Modelling this wearable-derived data offers a window into HR oscillations in real-world settings ^2–5^, describing the circadian autonomic modulation of cardiovascular physiology ^1^. As detected through photoplethysmography-equipped wearables, these oscillations have been associated with age, sex ^4,5^, risk factors for noncommunicable diseases ^4^, and pathological conditions, including heart failure ^2^, and clinical frailty ^3^. Such findings recently led the *American Heart Association* to list HR circadian rhythms among possible contributors to cardiometabolic health, expanding the boundaries of cardiometabolic risk evaluation and prevention ^6^. Despite such relevance in health outcomes, the correlates of real-world HR circadian rhythms remain scarcely characterized. Specifically, no literature investigated the relationship between HR circadian rhythms and basic lifestyle and chronobiological factors which longitudinally affect mortality risk and noncommunicable disease development, i.e., smoking, alcohol consumption^7–9^, multidimensional sleep health ^7,10^, and chronotype ^11^.

Within the broader context of sleep health, HR disruptions have been extensively characterized in chronic insomnia ^12,13^ - a disorder characterized by persistently disrupted sleep and concomitant daytime symptoms ^14^ that longitudinally increases cardiometabolic risk. In detail, nighttime HR alterations are currently considered as an autonomic hallmark of hyperarousal ^12^, a state of cognitive-emotional activation that impairs sleep and contributes to daytime symptoms in patients with insomnia^15,16^. Although modern conceptualizations define insomnia as a 24-hour disorder ^14^, previous research investigated HR mainly through nighttime polysomnographic studies ^12^, with no study performing a characterization of HR circadian rhythms through continuous, prolonged HR recordings in free-living settings ^12^.

Albeit wrist wearables equipped with photoplethysmography represent a valuable digital health tool to assess HR circadian oscillations in real-world settings, technical and methodological limitations may impair their reliability and hinder between-study comparability. Specifically, wearable-derived HR data shows strong agreement with prolonged electrocardiography (ECG) during sleep, resting wakefulness, and low-intensity physical activity ^1,17–21^. However, the accuracy of these wearable-derived HR measurements significantly decreases during moderate-to-vigorous exertion ^1,17–19^, mostly due to motion artifacts ^17,19^. This reduces HR data signal-to-noise ratio outside laboratories ^1,5,17,19^, which can cloud the interpretation and validity of research findings, especially when relatively small cohorts are studied ^1^. Strategies for handling this unreliability in continuous recordings vary from data rejection ^5^ to data fusion ^19^. Finally, the comparison of published literature on HR circadian rhythms is partially limited by methodological heterogeneity in the analysis of wearable-derived data. This heterogeneity results in different - but complementary - perspectives on HR circadian oscillations ^2–5^.

Employing multiple analytical methods of wearable-derived data, this study aimed at describing the impact of multiple cardiometabolic health determinants on real-world HR circadian rhythms in a sample of individuals without cardiometabolic diseases. To enhance the robustness of our results, we additionally developed a data-driven model enhancing the agreement between HR measurements from wearables and prolonged ECG monitoring.

## MATERIALS AND METHODS

### DATA AVAILABILITY STATEMENT

The data underlying this article will be shared on reasonable request to the corresponding author.

### ELIGIBILITY CRITERIA

Participants fulfilling the following inclusion criteria were recruited: assuming no antihypertensive drugs nor any other drug potentially affecting HR; suffering from no known neurologic, psychiatric, pulmonary and cardiometabolic disorders; fluency in Italian; not being an overnight shift worker; providing the signed informed consent. Healthy participants had no history of sleep disorders, while patients with insomnia presented with no other known sleep conditions. Chronic insomnia disorder was diagnosed based on the International Classification Sleep Disorders, 3^rd^ Ed. Text Revision ^14^.

### STUDY DESIGN, PROCEDURE, AND ETHICS STATEMENT

This study was structured into 2 phases. First, data from the cross-sectional study by Benedetti et al ^17^ was retrieved to develop and validate a statistical model to improve the accuracy of wearable-derived HR data (Phase 1). The study procedure is elsewhere extensively documented ^17^ and here summarized: in 2021, 25 healthy volunteers were consecutively recruited at the University of Pisa and concurrently monitored overnight through a Fitbit wearable device and a portable PSG (pPSG). Each participant spent ≥12-hr wearing both the wearable and the pPSG.

Second, a cross-sectional study was conducted between 2023 and 2025, by consecutively enrolling healthy community volunteers and patients with chronic insomnia without known cardiometabolic disorders (Phase 2). Patients with insomnia were recruited during their routine visits at the Sleep Medicine Clinic of the Pisa University Hospital, when they underwent a comprehensive clinical evaluation. After screening for eligibility criteria, participants were given both a wearable device to be continuously worn for a week, and a digital survey combining self-report questionnaires and basic questions on demographic and lifestyle data. The questionnaires explored participants’ sleep quality, sleepiness, and chronotype.

All participants provided an informed consent; the studies were conducted in compliance with the principles of the Declaration of Helsinki and were approved by the University of Pisa Bioethical Committee (Identifier 0036352/2020) and the Bioethical Committee of Calabria Region (Identifier 57/2025). Data has been reported according to the STROBE guidelines for cross-sectional studies ^22^.

### MEASURES

#### Self-report Questionnaires and Demographic, Lifestyle, and Clinical Data

The following demographic and lifestyle data were collected: age, height and weight, body mass index (BMI), sex, weekly cigarettes, and alcohol consumption. Alcohol consumption was recorded as weekly alcohol units (AU), with an AU corresponding to a half-pint of beer, a glass of wine, or a measure of spirit ^23^. All participants were of Caucasian ethnicity.

The following self-report questionnaires were employed to investigate sleep quality, sleepiness, and chronotype (Supplementary Material 1A for a detailed description): Pittsburgh Sleep Quality Index (PSQI) ^24^, Epworth Sleepiness Scale (ESS) ^25^, reduced Morningness-Eveningness Questionnaire (rMEQ) ^26^.

#### Wearable Devices

Two Fitbit wearables were employed. In Phase 1, a Fitbit ChargeHR^®^ was continuously worn on the non-dominant wrist during the monitoring period. In Phase 2, all subjects continuously wore a Fitbit Inspire 2^®^ for seven days on their non-dominant wrist. Both wearables are equipped with the PurePulse® light-emitting photoplethysmographic sensor (PPG) for HR detection ^17,27^, along with the same micro-electro-mechanical system triaxial sensor for accelerometry. A Fitbit proprietary algorithm employs accelerometric and HR data to estimate daily steps and min-level energy expenditure, as assessed by calories ^21^. Daily steps (thousands) and min-level calorimetric and HR data were obtained via an application programming interface provided by Fitbit through a third-party platform (www.sleepacta.com). Linear interpolation was employed to impute missing HR data, which represented 0.51% of the wearable-derived epochs and were caused by the PPG displacement.

#### Accelerometric Sleep Parameters

To obtain minute-level sleep-wake classification, calorimetric data were analysed through the *Dormi* algorithm by Sleepacta s.r.l., a validated and certified artificial neural network-based routine ^28^ registered as a medical, risk class I device within the European Database on Medical Devices (EUDAMED) (UDI-DI: PP13374SLEEP78/IFA). The sleep-wake classification obtained through the Dormi algorithm has been previously employed to estimate the following quantitative sleep parameters ^29,30^ across multiple clinical and healthy cohorts: total sleep time (TST), waking after sleep onset (WASO), sleep efficiency (SE), mid-sleep point, sleep regularity index (SRI). Sleep parameters definitions are available in Supplementary Material 1B.

#### The Sleep Health Index

A sleep health composite index was computed based on the RU-SATED framework, including 6 dimensions of sleep, based on their association with negative health outcomes: regularity, satisfaction, alertness, timing, efficiency, and duration ^31^. Based on previous studies ^32–34^, each dimension was operationalized using measurements derived from actigraphy or questionnaires and subsequently dichotomized (0 poor sleep, 1 good sleep), as detailed in Supplementary Material 1C. The scores from the six different sleep measures were summed, resulting in a 0-6 range. Higher scores indicate greater sleep health.

#### Portable PSG

The pPSG recording procedure performed in Phase 1 procedure is detailed in the work by Benedetti et al. ^17^ and here summarized. pPSG recordings were acquired using a MORPHEUS-HOME-LTM device (Micromed). Sleep scoring was conducted according to AASM guidelines ^35^. The electrocardiographic (ECG) signal was digitally processed to facilitate the detection of QRS events. The detection was performed automatically using MATLAB routines, and then manually verified. The HR was therefore calculated as the number of QRS complexes per minute, and aligned to the Fitbit-estimated HR.

### DATA ANALYSIS

Data were analyzed in RStudio 4.4.1. Mean (M) and standard deviation (SD) described quantitative variables, a table of frequencies and percentages presented categorical variables. To test possible differences between pairs of variables, parametric tests were employed. The significance level was set at p<0.05. Multiple analytical approaches were employed in this study, and here summarized.

The statistical plan employed in Phase 1 is detailed in Supplementary Material 2A. This procedure allowed for the development of a generalized additive model (GAM) designed to enhance the accuracy of wearables HR measurement in the absence of polysomnographic ECG data, which was subsequently applied to data gathered during Phase 2. The GAM was validated through a 10-fold cross-validation procedure and the mean percentage reduction in the root mean square error (RMSE) across validation folds was calculated to assess the reduction in HR estimation error, reflecting the improvement in the accuracy of wearable-derived HR measurement after data correction. After applying the GAM to raw HR time series gathered during Phase 2, the processed HR time series were analysed to describe HR circadian rhythms.

In Phase 2, functional-on-scalar (FoSR) modelling, nonparametric, and parametric analyses were employed to describe HR circadian rhythms and their relationships with the variables of interest, i.e., age, sex, lifestyle factors, sleep health, chronotype, and chronic insomnia diagnosis. While FoSR modelling provided a robust framework to visually assess time-locked associations between the independent predictors and HR circadian rhythms ^36^, parametric and nonparametric methods allowed for the quantification of the impact of such variables on circadian rhythms timing and robustness ^37,38^. Except for sex and chronic insomnia diagnosis, all the variables of interest were considered as continuous.

FoSR modelling was previously applied to wearable-derived accelerometric and HR data ^36^ and here implemented to obtain a visual representation of the relationship between the scalar independent predictors and HR as a function of time ^36^. Operationally, a 24hr average HR profile was first obtained for each participant by performing a min-level HR mean across all recording days ^36^. GAMs were subsequently fitted through the “bam” function (R package “mgcv”) ^39^. Specifically, GAMs were estimated by modelling the independent predictors as penalized cyclic cubic splines with a maximum of 7 bases and adding a random intercept to account for residual correlation ^36^. Predicted HR values and the independent predictors coefficients were finally plotted across the 24 hrs. Root mean square (RMS) effect sizes were estimated for independent predictors to compare their relative impact on HR circadian oscillations ^36^. The standardization procedure for numerical predictors was carried out by dividing each mean-centred variable by 2 SDs, ensuring comparability between binary predictors and numeric predictors ^40^.

Nonparametric and parametric analyses of prolonged time-series have been widely employed in rest-activity rhythms characterization and previously applied to describe wearable-derived HR circadian rhythms ^2,3^. To estimate parametric HR circadian rhythm parameters, i.e., Midline Estimating Statistic of Rhythm (MESOR), amplitude, R2, and acrophase ^41^, we applied the cosinor method by fitting a sine wave to HR data through the R package ‘Cosinor’ ^37^. Non-parametric measurements of HR rhythms, i.e., interdaily stability (IS), intradaily variability (IV), relative amplitude (RA), L5, M10, L5 start time, and M10 start time ^38^ were computed using the R package Non-Parametric Measures of Actigraphy Data ‘nparACT’ ^42^. The definition of each circadian parameter is provided in Supplementary Material 1D. Among circadian rhythm parameters, IV, relative amplitude and amplitude were considered as proxies of circadian robustness, while IS and R2 represented measures of circadian rhythm stability across the recording week.

Following HR circadian parameters estimation, multiple linear regression models were subsequently estimated to test age, sex, lifestyle variables, chronotype, sleep health, and chronic insomnia diagnosis as possible predictors of each HR circadian parameter. For each model, potential multicollinearity among the independent predictors was evaluated through the Variance Inflation Factor (VIF) ^43^.

After results inspection, we conducted a post-hoc receiver operating characteristic (ROC) curve analysis to evaluate the ability of the M10 start time to discriminate patients with insomnia from healthy controls, deriving an optimal threshold via the Youden method ^44^. Finally, to transparently illustrate the impact of our data processing pipeline and show how analysing raw data might affect the characterization of circadian rhythms, we replicated the analyses from Phase 2 on raw HR data (Supplementary Material 3).

## RESULTS

### PHASE 1: IMPROVING THE ACCURACY OF WEARABLE-DERIVED HR MEASUREMENT

A total of 10056 one-minute epochs of concurrent portable PSG (pPSG) and wearable-derived HR data were recorded from 25 healthy participants (mean age 22.4 years [SD 3], 68% females). First, HR data were visualized through a Bland-Altman Plot (Figure 1, panel A) and the limits of agreement from the Bland-Altman plot were subsequently employed to categorize epochs into 2 agreement groups. Specifically, the 445 epochs falling outside the limits of agreement were included in the low agreement group, whereas epochs remaining within these boundaries constituted the adequate agreement group.

**FIGURE 1:**
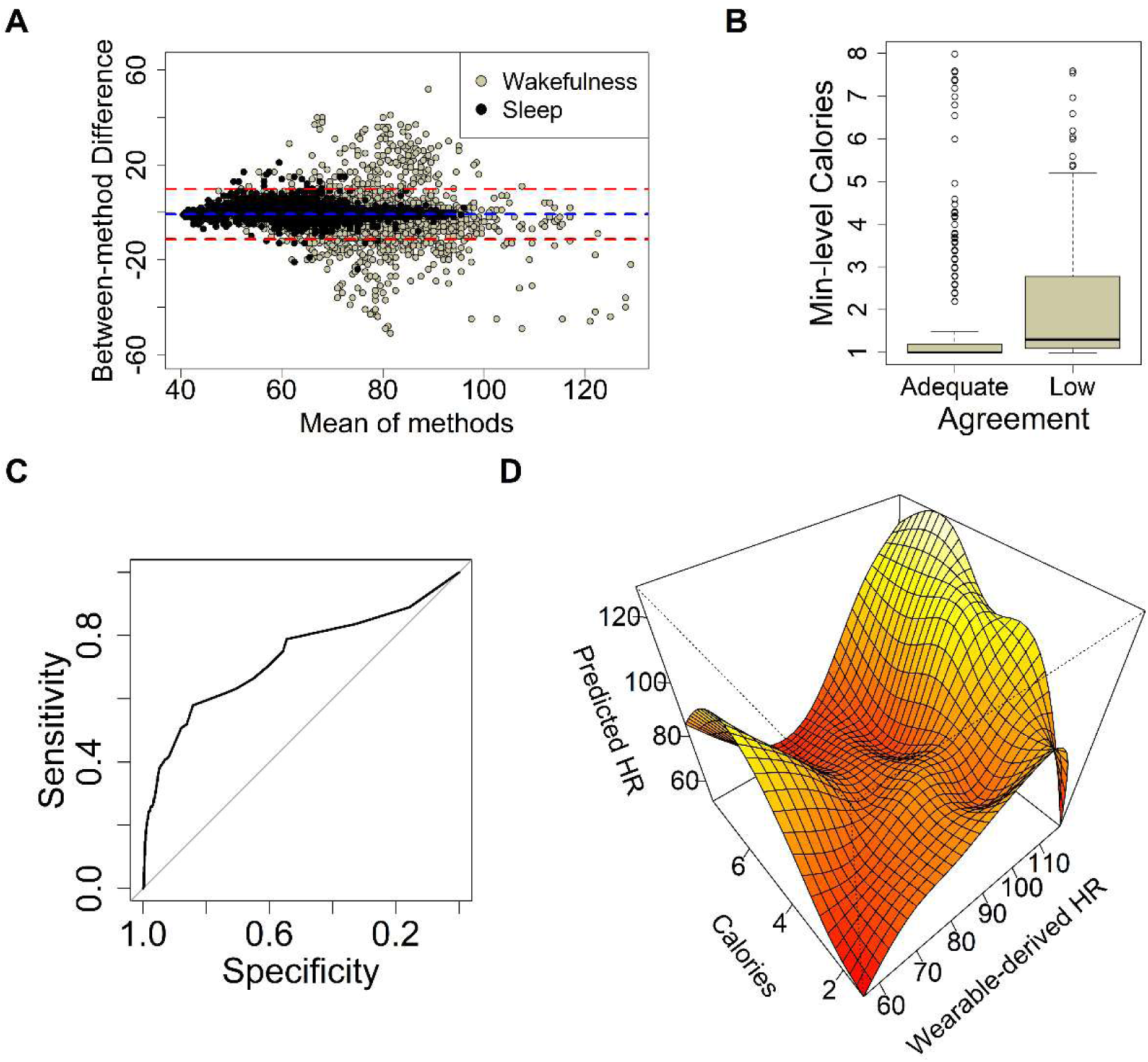
Development of a Generalized Additive Model to Improve the Accuracy of Wearable-derived Heart Rate Data. Phase 1 aimed at developing a statistical model to enhance the accuracy of wearable-derived hea1 rate (HR) data. Here, key steps from this process are represented. **Panel A:** A Bland-Altman plot representing the concordance between wearable-derived and pPSG HR measurements on 10,056 one-minute recording epochs. The plot displays the mean of the two methods against their absolute difference; horizontal lines indicate the mean bias and the limits of agreement (mean bias ±1.96 standard deviations); epochs outside the limits of agreement are chategorized as low agreement epochs; **Panel B:** During wakefulness, minute-level calories are significantly higher when the between-method agreement is low (Cohen’s d 1.25, p<0.001); **Panel C:** The receiver operating characteristic curve representing the ability of minute-level calories to discriminate between epochs of low and adequate agreement during wakefulness; **Panel D:** The predicted HR values from the validated generalized additive model are plotted against min-level calories and wearable-derived HR, showing the non-linear interaction between these independent predictors.

Aligning with previous literature ^1,18–21,45–47^, low agreement was robustly associated with wakefulness and physical activity. Specifically, the vast majority of epochs in the low agreement group were recorded when participants were awake (93.5%, 416 over 445 epochs; adjusted Cramer’s ϕ 0.19, p<0.001). Moreover, low HR estimation accuracy was strongly associated with increased physical activity during wakefulness, as assessed by calories (Cohen’s d 1.25, p<0.001; Figure 1, Panel B). Based on these data, a ROC curve was subsequently plotted to investigate the performance of min-level calories in distinguishing agreement groups during wakefulness (Figure 1, Panel C). A 1.243 calories threshold was derived, yielding an 85% specificity, a 58% sensitivity, and a 0.73 area under the curve (AUC).

When categorizing data based on such threshold during wakefulness, 972 1-minute epochs exceeded the calories threshold. Data from these epochs informed the 10-fold cross-validation of a GAM, which reduced the mean RMSE by 10.4% (paired Student’s t-test p<0.001). In Figure 1, Panel D, the predicted HR values from the validated GAM are plotted against min-level calories and wearable-derived HR, showing the non-linear interaction between the independent predictors. Specifically, the validated GAM performs an upward correction when higher caloric expenditure is recorded, especially at extreme HR values. Contrarily, high wearable-derived HR values are attenuated when caloric expenditure is low. This correction decreased the mean absolute error of wearable-derived HR from 8.3 bpm to 7.0 bpm during wakefulness epochs exceeding the 1.243-calories threshold. In Supplementary Material 2B, we provide further evidence that the validated GAM improves the accuracy of HR data in the low agreement group, with a negligible impact on accurate wearable-derived HR data.

### PHASE 2: WEARABLES TO ASSESS HR CIRCADIAN RHYTHMS IN HEALTHY PARTICIPANTS AND PATIENTS WITH CHRONIC INSOMNIA

Overall, 245 participants were recruited (Table 1). The GAM model developed in phase 1 was applied to wearable-derived HR time series, aiming at enhancing the accuracy of HR measurement and improving the robustness of the following results.

**TABLE 1.** Participants’ Data, Study 2 (n=245)

| TABLE 1 Participants' Data, Study 2 (n=245) |  |
| --- | --- |
| Variables | Mean (SD), n (%) |
| Age | 33 (17) |
| Sex (Female) | 129 (53%) |
| BMI | 23.0 (3.3) |
| Daily Steps (thousands) | 11.3 (4.4) |
| Weekly Alcohol Units | 2.15 (3.54) |
| Weekly Cigarettes | 7 (23) |
| rMEQ | 14.7 (3.9) |
| Sleep Health Index | 4 (1) |
| Chronic Insomnia Diagnosis | 31 (13%) |
| PSQI | 6.4 (4.0) |
| ESS | 5.6 (3.2) |
| Actigraphic TST | 6.91 (1.33) |
| Actigraphic WASO | 53 (42) |
| Actigraphic SE | 89 (10) |
| Actigraphic SRI | 72 (16) |
| HR Circadian Parameters |  |
| Acrophase | 16.20 (2.19) |
| Amplitude | 10.0 (3.2) |
| MESOR | 73.5 (5.3) |
| Variables | Mean (SD), n (%) |
| R2 | 0.54 (0.16) |
| IS | 0.59 (0.15) |
| IV | 0.57 (0.18) |
| RA | 0.14 (0.05) |
| L5 | 61 (8) |
| M10 | 80.6 (4.8) |
| L5 Start Time | 2.08 (1.44) |
| M10 Start Time | 11.26 (2.91) |

BMI: Body Mass Index (kg/m<sup>2</sup>), PSQI: Pittsburgh Sleep Quality Index; rMEQ: reduced Morningness-Eveningness Questionnaire; ESS: Epworth Sleepiness Scale; TST: Total Sleep Time (hours); WASO: Wake After Sleep Onset (minutes); SE: Sleep Efficiency (%); Sleep Regularity Index (%); SD: standard deviation; the Sleep Health Index ranges between 0 and 6, with higher scores suggesting healthier sleep. MESOR: Midline-Estimating Statistic Of Rhythm (bpm); IS: Interdaily Stability; IV: Interdaily Variability; RA: Relative Amplitude; M10: the average HR during the 10-hour period of peak HR across the day (bpm); L5: the average HR during the 5-hour period of nadir HR across the day (bpm).

A FoSR model was subsequently estimated to visualize the HR circadian rhythms and investigate the impact of age, sex, lifestyle variables, sleep health, chronotype, and chronic insomnia diagnosis on HR as a function of time. The predicted HR values and the time-varying coefficients of the independent predictors (95% confidence intervals) are represented in Figure 2 and 3, respectively. Moreover, HR circadian parameters and the linear regression models investigating the impact of the aforementioned predictors are presented in Table 1 and Table 2, respectively. No multicollinearity was detected in these models after VIF estimations.

**FIGURE 2:**
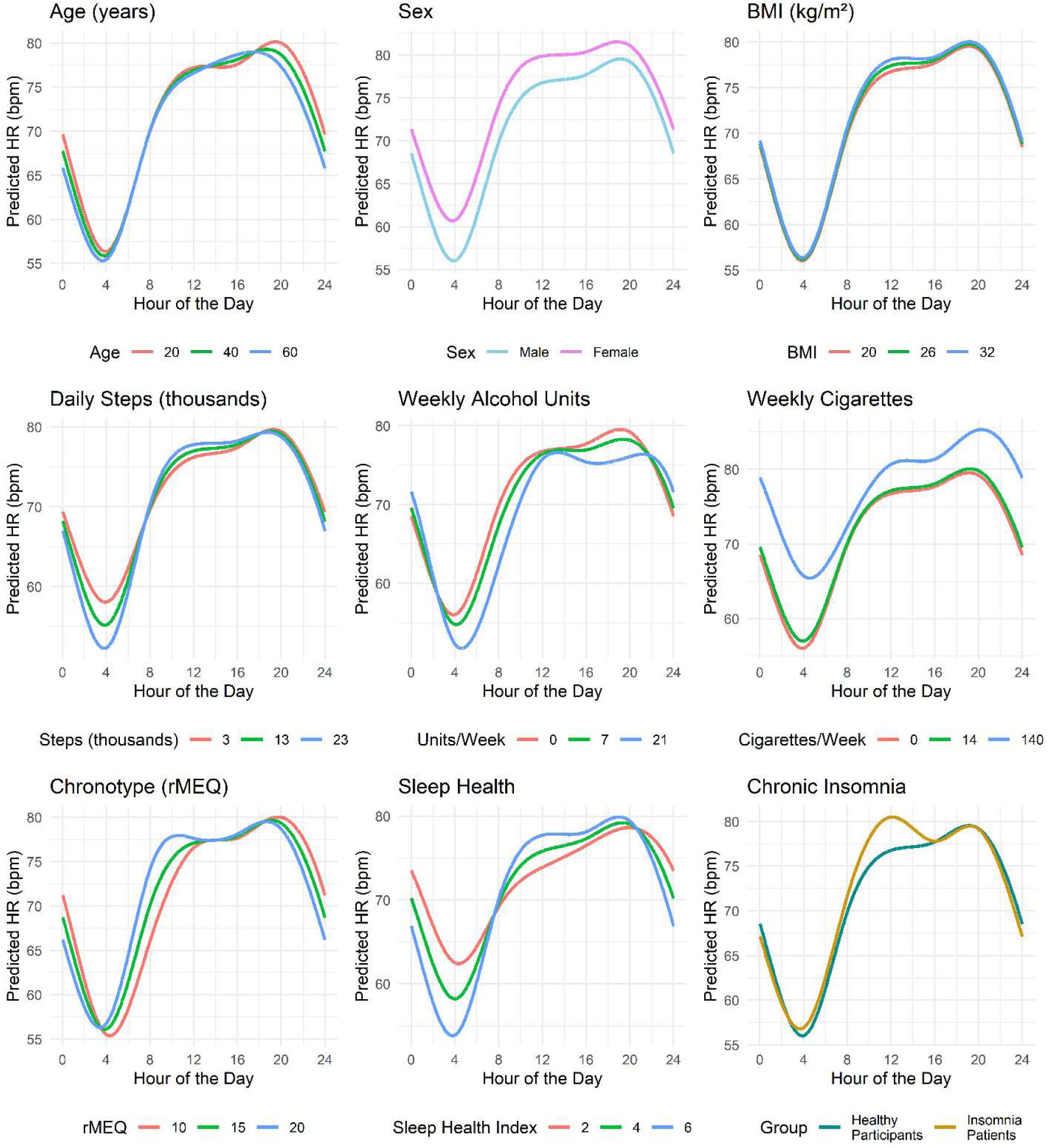
Heart Rate Circadian Rhythms as a Function of Demographics, Lifestyle, Sleep Health, Chronotype, and Chronic Insomnia. A function-on-regression (FoSR) model was estimated to visualize time-locked associations between heart rate (HR) and the included independent predictors. In this figure, each plot represents predicted HR values obtained by varying one independent predictor and fixing others at reference values. Reference values: Age= 30; Sex= Male; body-mass index (BMI) = 20; Daily Steps= 10.000; Sleep Health Index = 5; reduced Morningness-Eveningness Scale (rMEQ) = 15, Weekly Cigarettes = 0, Weekly Alcohol Units= 0, Insomnia Diagnosis: patients without insomnia. Sleep Health ranges between 0 and 6, with higher values suggesting healthier sleep; higher rMEQ scores indicate higher tendencies towards morningness; an alcoholic unit corresponds to a half-pint of beer, a glass of wine, or a measure of spirit

**FIGURE 3:**
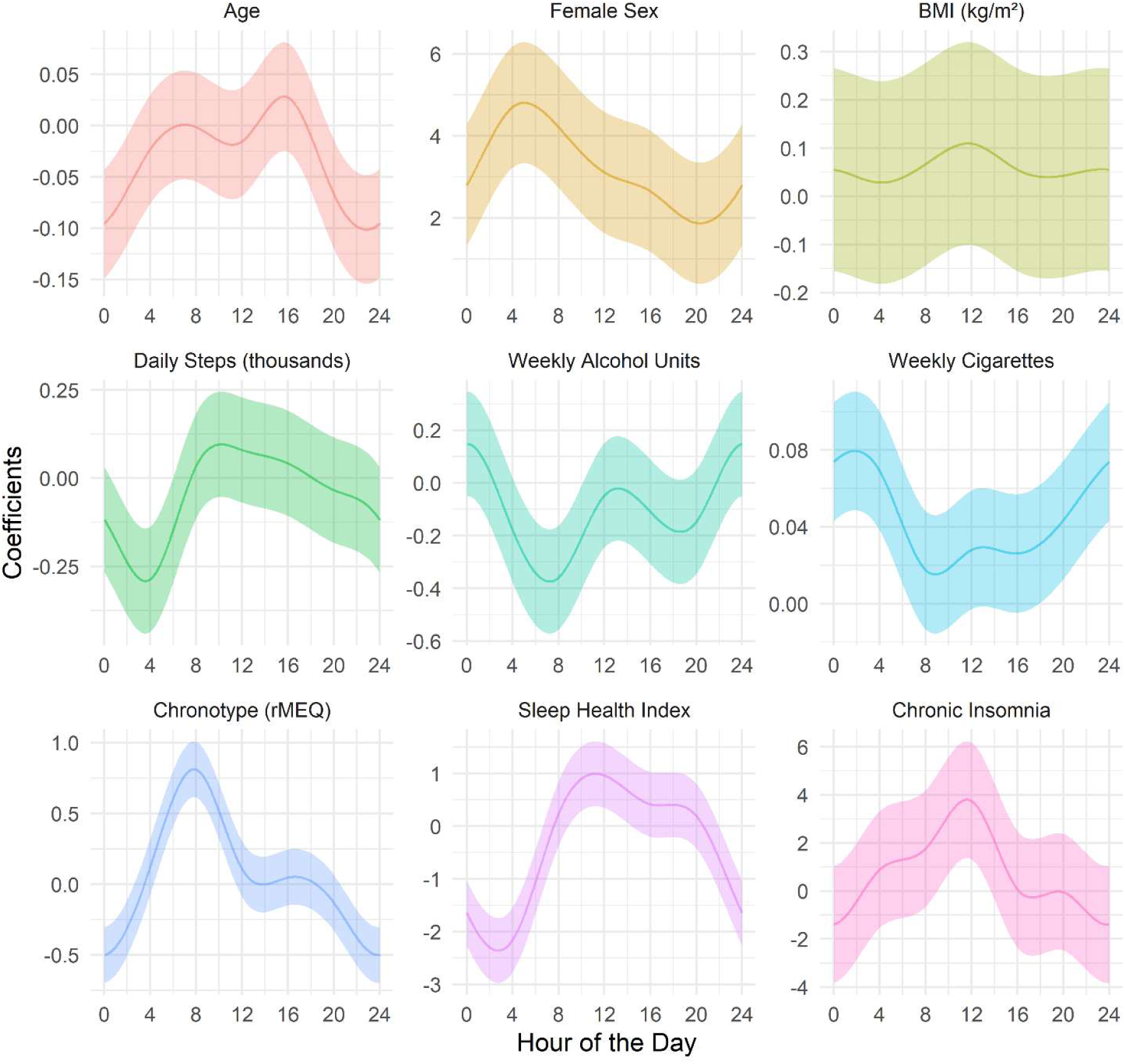
Time-varying Coefficients with 95% Confidence Intervals of Heart Rate Independent Predictors. A function-on-regression (FoSR) model was estimated to visualize lime-locked associations between heart rate (HR) and the included independent predictors. In this figure, the time-varying coefficients of HR independent predictors are represented along with with their 95% confidence intervals (Cls). An independent predictor is statistically significant at any given time point when the 95% Cl of its coefficient lies entirely above or below the zero threshold. The predicted coefficiens are obtained by varying one independent variable in the FoSR model and fixing others at reference values. Reference Values Age= 30; Sex= Male; body-mass index (BMI) = 20; Daily Steps = 10.000; Sleep Health Index= 5; reduced Morningness-Eveningness Scale (rMEQ) = 15, Weekly Cigarettes = 0, Weekly Alcohol Units = 0, Insomnia Diagnosis: patients without insomnia. Sleep Health ranges between 0 and 6, with higher values suggesting healthier sleep; higher rMEQ scores indicate higher tendencies towards morningness; an alcoholic unit corresponds to a half-pint of beer, a glass of wine, or a measure of spirit

**TABLE 2:** Multiple Linear Regression Models assessing Independent Predictors of Heart Rate Circadian Parameters.

| **Covariates** | **Acrophase** | **Amplitude** | **MESOR** | **R2** | **IS** | **IV** | **RA** | **L5** | **M10** | **L5 Start Time** | **M10 Start Time** |
| --- | --- | --- | --- | --- | --- | --- | --- | --- | --- | --- | --- |
| **Age** | -0.004<br>(0.68) | 0.01<br>(0.41) | -0.03<br>(0.28) | 0.003<br>**(0.001)** | 0.003<br>**(<0.001)** | -0.001<br>(0.32) | 0<br>(0.80) | -0.02<br>(0.62) | -0.02<br>(0.33) | -0.002<br>(0.74) | -0.01<br>(0.52) |
| **Female Sex** | -0.54<br>(0.053) | -1.26<br>**(0.004)** | 3.19<br>**(<0.001)** | -0.03<br>(0.26) | 0<br>(>0.9) | 0.07<br>**(0.013)** | -0.02<br>**(<0.001)** | 4.7<br>**(<0.001)** | 2.33<br>**(<0.001)** | -0.06<br>(0.73) | -0.57<br>(0.13) |
| **BMI** | -0.01<br>(0.77) | -0.04<br>(0.48) | 0.05<br>(0.62) | 0.001<br>(0.85) | 0.001<br>(0.85) | 0<br>(0.92) | -0.001<br>(0.42) | 0.11<br>(0.47) | 0.03<br>(0.75) | -0.01<br>(0.70) | -0.03<br>(0.59) |
| **Daily Steps** | -0.07<br>**(0.020)** | 0.20<br>**(<0.001)** | -0.05<br>(0.53) | 0<br>(0.88) | 0.01<br>**(0.005)** | -0.001<br>(0.60) | 0.003<br>**(<0.001)** | -0.31<br>**(0.004)** | 0.13<br>(0.059) | -0.02<br>(0.22) | -0.06<br>(0.11) |
| **Weekly AUs** | 0.02<br>(0.59) | 0.01<br>(0.84) | -0.12<br>(0.24) | 0<br>(0.90) | -0.001<br>(0.68) | 0<br>(0.90) | 0.001<br>(0.54) | -0.2<br>(0.22) | -0.14<br>(0.12) | 0.03<br>(0.17) | 0.02<br>(0.64) |
| **Weekly Cigarettes** | 0.013<br>**(0.025)** | -0.01<br>(0.18) | 0.04<br>**(0.006)** | 0<br>(0.62) | -0.001<br>**(0.041)** | 0<br>(0.77) | 0<br>**(0.042)** | 0.06<br>**(0.005)** | 0.03<br>**(0.025)** | 0.009<br>**(0.022)** | 0.015<br>(0.06) |
| **rMEQ** | -0.2<br>**(<0.001)** | -0.11<br>(0.053) | 0.07<br>(0.454) | -0.01<br>(0.053) | 0.002<br>(0.37) | 0.002<br>(0.51) | -0.001<br>(0.19) | 0.16<br>(0.26) | 0.02<br>(0.81) | -0.17<br>**(<0.001)** | -0.23<br>**(<0.001)** |
| **Sleep Health Index** | -0.348<br>**(0.003)** | 0.886<br>**(<0.001)** | -0.338<br>(0.28) | 0.032<br>**(<0.001)** | 0.031<br>**(<0.001)** | -0.011<br>(0.32) | 0.011<br>**(<0.001)** | -1.183<br>**(0.007)** | 0.216<br>(0.45) | -0.115<br>(0.13) | -0.41<br>**(0.009)** |
| **Insomnia** | -1.04<br>**(0.024)** | 0.63<br>(0.38) | 0.85<br>(0.49) | 0.06<br>(0.12) | 0.04<br>(0.27) | -0.05<br>(0.26) | 0.001<br>(0.92) | 0.84<br>(0.62) | 1.55<br>(0.17) | -0.03<br>(0.92) | -2.18<br>**(<0.001)** |
| **R2** | 0.30 | 0.23 | 0.12 | 0.12 | 0.20 | 0.06 | 0.25 | 0.20 | 0.08 | 0.31 | 0.28 |
| **Adjusted R2** | 0.28 | 0.21 | 0.08 | 0.09 | 0.17 | 0.02 | 0.22 | 0.17 | 0.05 | 0.28 | 0.25 |

β coefficients and p values are showed for each independent predictor. In bold: p<0.05
Higher rMEQ scores indicate increased morningness. A higher Sleep Health Index suggests healthier sleep (0-6 range). Variables encoding temporal data are expressed as hours after midnight.
AUs: Alcohol Units; BMI: body-mass index (kg/m<sup>2</sup>); rMEQ: reduced Morningness-Eveningness Scale; MESOR: Midline-Estimating Statistic Of Rhythm (bpm); IS: Interdaily Stability; IV: Interdaily Variability; RA: Relative Amplitude; M10: the average HR during the 10-hour period of peak HR across the day (bpm); L5: the average HR during the 5-hour period of nadir HR across the day (bpm).

Older participants showed more stable HR circadian rhythms as assessed by R2 (β=0.003; p=0.001) and IS (β=0.003; p<0.001), with significantly lower HR between 19:29 and 02:27. Sex-effect analyses highlighted higher HR values across the 24 hours in females, with an average 3.19-bpm increase as assessed by MESOR (p<0.001). However, the sex disparities were more pronounced during the circadian nadir (L5 β=4.7 bpm; p<0.001) compared to daytime (M10 β=2.33 bpm; p<0.001), peaking at about 05:00. Finally, females showed significantly lower robustness of HR circadian rhythms, as assessed by amplitude (β=-1.26, p=0.004), RA (β=-0.024, p<0.001), and IV (β=0.067; p=0.013). Notably, BMI did not predict HR circadian rhythms variations. Participants performing more daily steps experienced increased robustness and stability of HR circadian rhythms, as assessed by amplitude (β=0.20; p<0.001), RA (β=0.003; p<0.001), and IS (β=0.006; p=0.005), along with lower HR at night (00:34-05:58). Weekly AUs significantly predicted an early morning HR decrease (04:11-10:06), while increased smoking intensity predicted higher mean HR (MESOR β=0.043; p=0.006), with this effect being more pronounced between evening and early morning (17:55-06:42; L5 β=0.061, p=0.005; M10 β=0.033, p=0.025). Notably, smoking intensity also predicted lower stability of HR circadian rhythms (IS β=-0.001; p=0.041) and a delay in HR circadian timings (acrophase β=0.013, p=0.025; L5 start time β=0.009, p=0.022). A clear shift in HR circadian rhythms timing was observed across the morningness-eveningness spectrum, as reflected by both FoSR modelling and circadian rhythm parameters. Sleep health significantly impacted heart rate at night (21:59-06:39), with an average 1.18-bpm L5 increase for each impaired sleep health dimension (p<0.007). Moreover, better sleep health predicted higher circadian rhythm robustness and stability (amplitude β=0.89, p<0.001; RA β=0.01, p<0.001; R2 β=0.03, p<0.001; IS β=0.03, p<0.001). Notably, while impaired sleep health predicted delayed HR circadian rhythm timings (acrophase β=-0.35 hours, p=0.003; M10 start time β=-0.41 hours, p=0.009), patients with chronic insomnia showed an earlier HR morning peak (09:02 and 13:41; M10 start time β=-2.18 hours, p<0.001; β=-1.04 hours, p=0.024). Post-hoc ROC analysis found an optimal M10 start time threshold of 09:19 to distinguish patients with insomnia from healthy participants, yielding an 80% specificity and a 74% sensitivity.

When ranking the independent predictors based on their RMS standardized effect sizes, sex (3.33 bpm), chronotype (3.17 bpm), and sleep health (2.83 bpm), predominantly impacted HR circadian oscillations, followed by smoking intensity (2.28bpm), chronic insomnia diagnosis (1.86 bpm), age (1.68 bpm), weekly alcohol units (1.35 bpm), daily steps (1.22 bpm), and BMI (0.46 bpm).

## DISCUSSION

To the best of our knowledge, this is the first study to investigate HR circadian rhythms in individuals without cardiometabolic disorders during free-living conditions and by means of multiple analytical approaches. In detail, we described circadian HR oscillations across various demographic, lifestyle, and sleep-related conditions. Notably, this characterization highlighted sex, sleep health, and chronotype as pivotal modulators of HR circadian oscillations ^6,7,11,31^. Moreover, we employed a novel approach of prolonged, continuous monitoring in free-living conditions to explore HR circadian rhythms in insomnia, identifying previously unobserved insomnia markers. Notably, the robustness of our results is strengthened by the development of a model enhancing the agreement between wearable and ECG measurements of HR. Finally, modelling wearable-derived HR data by means of multiple analytical approaches provided a more comprehensive understanding of HR circadian rhythms and their correlates compared to previous literature relying on single methodologies.

Building upon the reported methodological framework, this study replicated previous results describing the relationships between HR circadian rhythms and basic demographic and lifestyle factors ^4,5^. In detail, we observed a reduced variability of HR circadian oscillations in older participants ^5^. Moreover, we confirmed that females experience higher mean HR across the 24 hours ^5^, with the sex-effect being more pronounced during the HR circadian nadir (+4.7 bpm) compared to daytime (+2.3 bpm) ^48^. This time-locked increase in sex differences may be primarily attributable to the sleep-related reduction in physical activity. Such reduction indeed minimizes the inter-individual HR variability dependent on physical activity, consequently fostering the expression of truly time-invariant HR differences ^48^. Consistent with a previous large-scale, real-world study ^5^, we additionally observed no differences in HR circadian rhythms related to BMI, suggesting that the effect of BMI on HR may be small in free-living conditions, especially when controlling for demographic and lifestyle factors. Finally, while corroborating the previously documented association between increased physical exertion and a shift of HR circadian oscillations towards earlier times, i.e., a phase advance ^4,5^, this study also provides the characterization of a reduced nocturnal HR nadir and a concomitant increase in HR circadian amplitude and stability linked to elevated physical activity in free-living conditions. Unlike previous research that omitted HR data recorded during physical exertion ^5^ or sleep periods ^4^, our use of uninterrupted monitoring captured the full physiological trajectory of HR oscillations. This is critical because daytime physical activity modulates subsequent sleep architecture; by increasing slow-wave sleep representation, it induces a deeper nocturnal HR nadir via enhanced parasympathetic activity—an effect that remains invisible when data collection is non-continuous ^49^. Beyond physical activity, chronotype and multidimensional sleep health represent modifiable lifestyle-related risk factors for negative cardiometabolic outcomes ^7,11,31^. The mechanisms mediating such relationship remain partially unknown, with previous literature hypothesizing impaired autonomic regulation of the cardiovascular system as a possible pathogenic pathway ^6,7,11^. This hypothesis is consistent with our results, which highlighted chronotype and multidimensional sleep health as pivotal modulators of HR circadian rhythms. In detail, we found chronotype to robustly predict HR circadian phase, indicating that the autonomic modulation of cardiovascular function is tightly aligned with circadian typology. Moreover, our findings indicate that lower sleep health is associated with a phase delay and reduced robustness and stability of HR circadian rhythms, as assessed by reduced amplitude of HR oscillations, increased day-to-day HR variability, and higher HR values during the circadian nadir. While prior literature has characterized nighttime sympathetic elevation in poor sleepers ^13^, our use of longitudinal 24-hour monitoring bridges a critical gap by identifying the daytime manifestations of these sleep-induced HR disruptions. These data demonstrate that the autonomic impact of poor sleep health persists well beyond the nocturnal period and reinforce the growing consensus that sleep and circadian health may represent main determinants of cardiometabolic outcomes ^6,7,11,31^.

In the recruited sample, individuals with chronic insomnia exhibited a significant phase advance in the morning peak of heart rate (HR), a finding that diverges from the HR circadian patterns associated with poor sleep health. This apparent inconsistency may stem from our statistical approach. Indeed, by simultaneously adjusting for both “sleep health” and “clinical insomnia diagnosis,” the “sleep health” metric effectively captured the non-specific variance associated with sleep duration, efficiency, regularity, satisfaction, timing and sleepiness. This statistical isolation consequently allowed for the identification of the unique pathophysiological impact that chronic insomnia exerts on diurnal cardiovascular rhythms, independent of broader sleep health metrics. This interpretation is supported by the discriminatory power of the 09:19 M10 start time threshold, yielding an 80% specificity and a 74% sensitivity in distinguishing healthy subjects from patients with insomnia. Moreover, this result aligns with modern conceptualizations of chronic insomnia as a 24-hour disorder ^14^, distinguishing insomnia-specific, daytime HR disruptions from sleep-related HR variations, which characterize multiple sleep disorders ^12,13^. These preliminary, hypothesis-generating findings highlight HR circadian phase as a possible marker of insomnia disorder in real-world settings, possibly mirroring the underlying degree of hyperarousal and peripheral sympathetic activity ^15^.

Finally, our study is the first to investigate the impact of smoke and alcohol consumption on HR circadian rhythms. These modifiable lifestyle-related behaviours represent leading risk factors for mortality and noncommunicable diseases ^7^, with the related autonomic modulations likely contributing to their impact on health outcomes ^8,9^. Notably, these autonomic modulations were characterized by dose-dependent, time-locked oscillations in HR. Specifically, smoking intensity was a significant predictor of elevated mean HR, with the most pronounced effects occurring during the nocturnal period (evening to early morning). This elevation resulted in a diminished circadian amplitude and a significant phase delay of the HR rhythm, consistent with the known sympathomimetic effects of smoking ^9^ and with the previously documented preference for eveningness among smokers ^32^. In contrast, higher alcohol consumption predicted a decrease in early-morning HR, aligning with the previously observed parasympathetic rebound following alcohol consumption ^8^.

Although the limited number of recruited patients with insomnia represents a primary limitation of this study, the extended duration of wearable monitoring strengthens our results. Furthermore, focusing on overall healthy individuals provides a valuable normative reference but limits the generalizability to pathological cases where risk factors may more drastically impact heart rate.

In summary, by establishing a methodological framework for the analysis of circadian HR rhythmicity via wearables, this study provides a foundation for more detailed autonomic phenotyping of cardiometabolic risk factors. Moreover, our findings suggest that characterizing these rhythms under free-living conditions may facilitate the detection of physiological hyperarousal in individuals with sleep-related pathologies. These data indicate that moving beyond static measures, such as resting heart rate, toward longitudinal chronobiological markers could offer a better understanding of cardiovascular homeostasis. Further prospective research is required to validate the predictive value of these dynamic markers against established clinical endpoints.

## ACKNOWLEDGMENTS

None.

